# The oral microbiome and all-cause mortality in the US population

**DOI:** 10.1101/2024.12.03.24318413

**Authors:** Emily Vogtmann, Yukiko Yano, Jianxin Shi, Yunhu Wan, Vaishnavi Purandare, Jody McLean, Shilan Li, Rob Knight, Lisa Kahle, Autumn G. Hullings, Xing Hua, Barry I. Graubard, Maura L. Gillison, J. Gregory Caporaso, Nicholas A. Bokulich, Martin J. Blaser, Neal D. Freedman, Anil K. Chaturvedi, Christian C. Abnet

**Affiliations:** Division of Cancer Epidemiology and Genetics, National Cancer Institute, Bethesda, MD, USA; Frederick National Laboratory for Cancer Research/Leidos Biomedical Research Laboratory, Inc, Frederick, MD, USA; National Center for Health Statistics, Centers for Disease Control and Prevention, Hyattsville, MD, USA; Johns Hopkins University, Baltimore, MD, USA; Center for Microbiome Innovation, University of California San Diego, La Jolla, CA, USA; Information Management Services, Inc, Calverton, MD, USA; Department of Nutrition, University of North Carolina, Chapel Hill, NC, USA; Department of Thoracic and Head and Neck Medical Oncology, MD Anderson Cancer Center, Houston, TX, USA; Center for Applied Microbiome Science, Pathogen and Microbiome Institute, Northern Arizona University, Flagstaff, AZ, USA; Laboratory of Food Systems Biotechnology, Department of Health Sciences and Technology, ETH Zurich, Zurich, Switzerland; Center for Advanced Biotechnology and Medicine, Rutgers, Piscataway, NJ, USA; Division of Cancer Control and Population Sciences, National Cancer Institute, Bethesda, MD, USA

**Author notes:** These authors contributed equally (Emily Vogtmann, Yukiko Yano, Anil K. Chaturvedi, Christian C. Abnet). **Correspondence** Christian C. Abnet, PhD, MPH, Director, Metabolic Epidemiology Branch, Division of Cancer Epidemiology & Genetics, National Cancer Institute 9609 Medical Center Drive, 6E410, Rockville, MD 20852.

## Abstract

**Importance:** Poor oral health, including periodontal disease, is associated with oral microbiome changes and increased mortality risk. However, no large studies have evaluated whether the oral microbiome is directly associated with mortality.

**Objective:** To evaluate whether measures of the oral microbiome is prospectively associated with all-cause mortality.

**Design:** A cross-sectional survey with samples collected from 2009-2012 and mortality linkage to the restricted-use National Death Index (NDI) through 2019.

**Setting:** The National Health and Nutrition Examination Survey (NHANES) 2009-2012, a multistage probability sample of the US population.

**Participants:** NHANES participants 20- to 69-years-old who were eligible for linkage to the NDI and provided oral rinse specimens (N=7,721, representing approximately 194 million individuals).

**Exposure:** Oral microbiome ascertained by sequencing the V4 region of the 16S rRNA gene of extracted DNA from oral rinse specimens. Alpha diversity, beta diversity, and genus-level data were generated using DADA2 and QIIME.

**Main outcome and measure:** All-cause mortality.

**Results:** After an average of 8.8 years, a total of 426 participants died. Using Cox proportional hazards regression and after controlling for multiple comparisons where appropriate, continuous alpha diversity was inversely associated with all-cause mortality, but only the association for the Shannon-Weiner index was significant with full adjustment for major risk factors (hazard ratio [HR] per standard deviation [SD]=0.85; 95% confidence interval [CI]=0.74-0.98). The principal coordinate analysis (PCoA) vector 2 from the Bray-Curtis dissimilarity matrix (HR per SD=0.83; 95% CI=0.73-0.93) and PCoA1 from weighted UniFrac (HR per SD=0.86; 95% CI=0.75-0.98) were significantly associated with all-cause mortality after full adjustment. Few associations were observed at the genus-level after Bonferroni correction, but an increase in 1 SD of the relative abundance of *Granulicatella* and *Lactobacillus* were associated with a 17% (95% CI=1.11-1.24) and 11% (95% CI=1.06-1.16) increase in mortality risk, respectively. Compared to participants with no detectable *Bacteroides*, participants in the highest tertile of *Bacteroides* had decreased mortality risk (HR=0.54; 95% CI=0.40-0.74).

**Conclusions and relevance:** Some measures of the oral microbiome were associated with all-cause mortality in this representative population cohort. These results suggest that oral bacterial communities may be important contributors to health and disease.

**Key points:** Question: Does the human oral microbiome impact an individual’s risk of mortality?

Findings: In this prospective study including 7,721 individuals of which 426 died over follow-up, specific measures of the oral microbiome were associated with all-cause mortality.

Meaning: The microbes living in the oral cavity may play an important role in human health.

## Introduction

The oral microbiome serves multiple functions in human health, locally and systemically. Specific oral microbes are closely linked to periodontal disease,^1^ and many others are associated with oral health conditions such as caries.^2^ The oral microbiome also has been associated with cardiovascular disease^3^ and multiple site-specific cancers including lung cancer.^4^ These microorganisms also play a central role in the metabolism of ethanol to its carcinogenic intermediate acetaldehyde.^5^ The oral microbiome may be related to mortality, as previous studies have identified associations between oral health conditions and mortality,^6,7^ but no large studies of the oral microbiome including genus-level taxonomy have been conducted to directly investigate mortality associations.

We analyzed associations between the oral microbiome from participants aged 20-69 years in the 2009-2012 cycles of the National Health and Nutrition Examination Survey (NHANES)^8^ and all-cause mortality from linkage to the National Death Index (NDI) while controlling for numerous potential confounders.

## Methods

NHANES is a continuous survey conducted by the National Center for Health Statistics (NCHS) to characterize the health and nutritional status of the US population. Participants are interviewed about demographics, diet, and other health-related data and then visit a mobile examination center (MEC) for biospecimen collection and health examinations.^9^ NDI mortality data through December 31, 2019 were available for eligible adult participants from the NCHS restricted-use linked mortality file.^10^

From 2009-2012, oral rinse samples were collected from participants aged 14-69 years which were used for microbiome analyses. After DNA extraction,^11^ the V4 region of the 16S rRNA gene was PCR amplified and sequenced. Sequencing data were processed using DADA2 and QIIME and taxonomy was assigned using the SILVA v123 database. After removal of the amplicon sequencing variant (ASV) SV1032, a human pseudogene, alpha and beta diversity metrics were generated at an even sampling depth of 10,000 reads per sample.^12^

As fully described in the companion manuscript (MEDRXIV/2024/318415), we incorporated many potential confounders in our analysis, including continuous age and spline variables for non-linear age relationships, sex, self-identified race and ethnicity categories grouped by NHANES standard practice, education, marital status, and the income-to-poverty ratio. Detailed categories of self-reported lifetime cigarette smoking history and alcohol consumption were used. Periodontal disease status, tooth counts, and body mass index categories were calculated based on health examinations. We included self-reported use of prescription medications, including antibiotics, anti-lipidemic drugs, inhaled respiratory drugs, and drugs for treatment of gastroesophageal reflux disease. Finally, we determined each participant’s diabetes and hypertension status using self-report, laboratory and examination data.

Analyses were restricted to MEC participants, aged 20-69 years with microbiome and NDI data (N=7,721, representing approximately 194 million people). We presented the characteristics of the population by mortality status and estimated hazard ratios (HR) and 95% confidence intervals (CI) for these characteristics from multivariable Cox proportional hazards models with time since the MEC visit as the underlying time metric. We estimated HRs for the association between the alpha diversity metrics (observed ASVs, Faith’s phylogenetic diversity [PD], Shannon-Weiner index, and Simpson index) and principal coordinate analysis (PCoA) vectors from the beta diversity matrices (unweighted UniFrac, weighted UniFrac, and Bray-Curtis). We estimated HRs for genus-level presence, relative abundance, centered log-ratio (CLR) transformed abundances, and a combined variable incorporating both the presence and tertiles of relative abundance. Bonferroni-adjusted p-values cutpoints were determined for genus-level models. We used SAS and SUDAAN and incorporated the complex survey design, including post-stratification weights.

## Results

After an average of 8.8 years of follow-up, a total of 426 participants died, including 135 deaths from malignant neoplasms and 91 from heart diseases. Participants who died were more likely to be older, male, experiencing poverty, categorized with Class 2 or 3 obesity, and have periodontal disease or edentulism (Table 1).

**Table 1.**
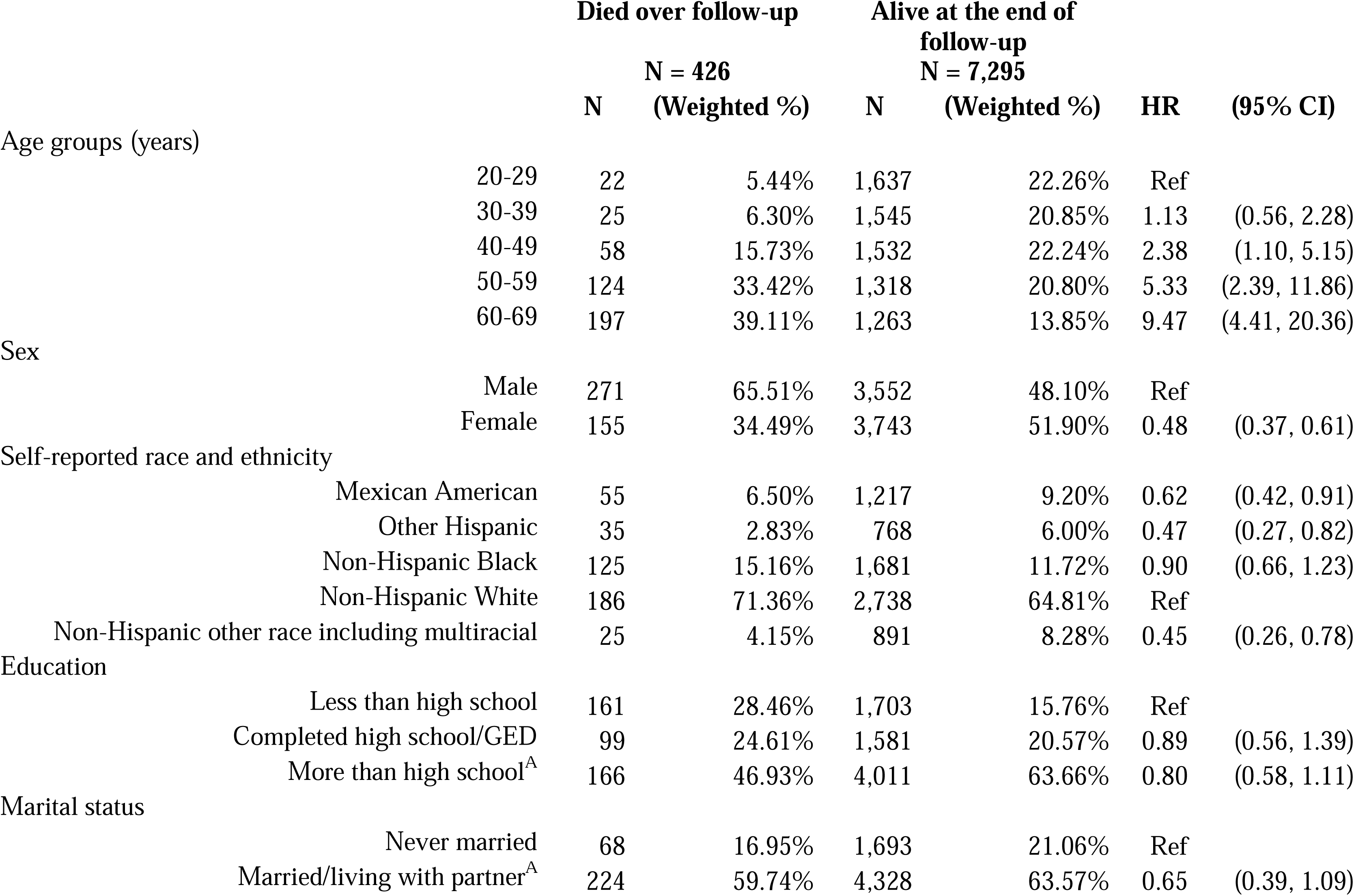
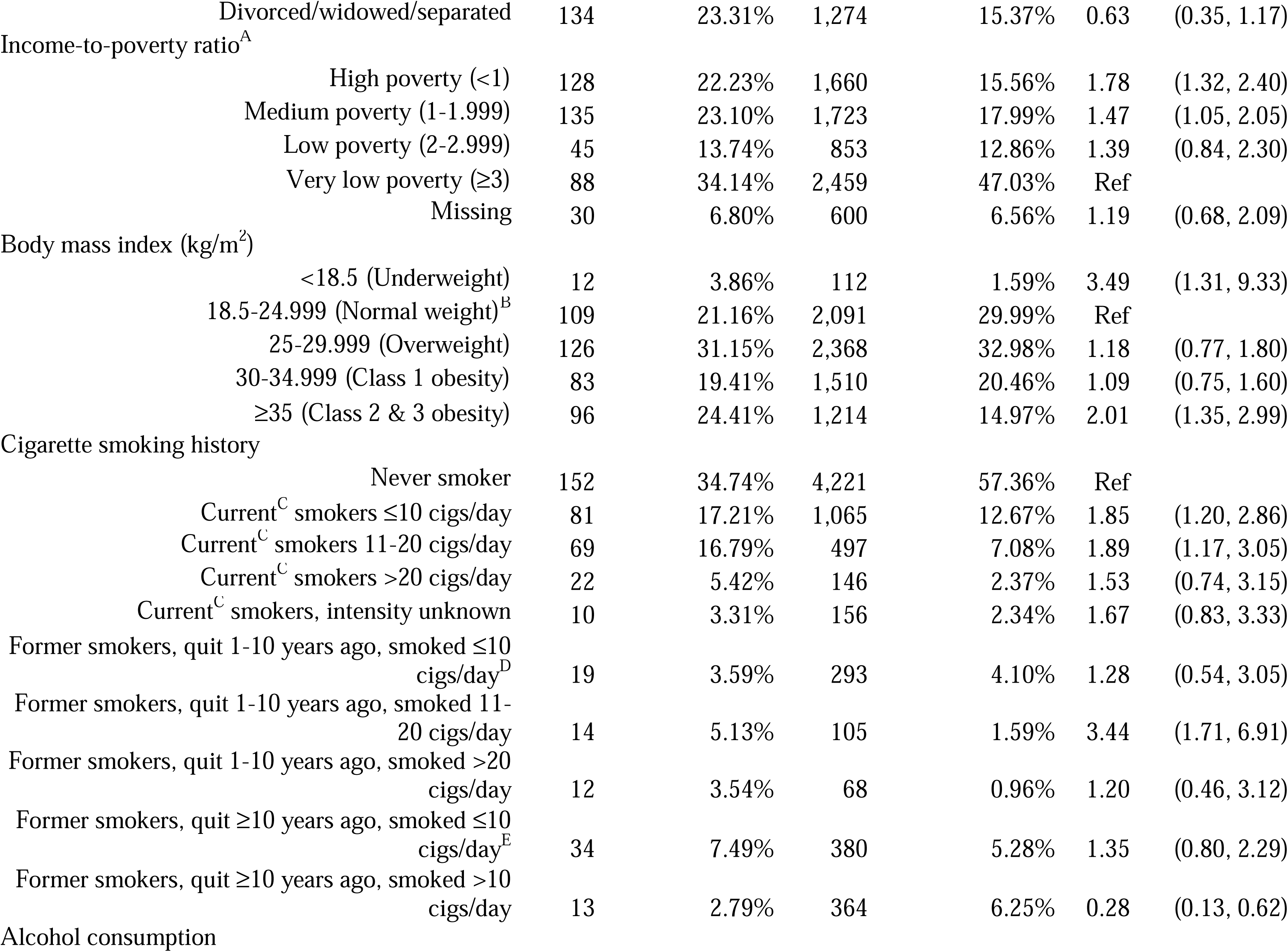
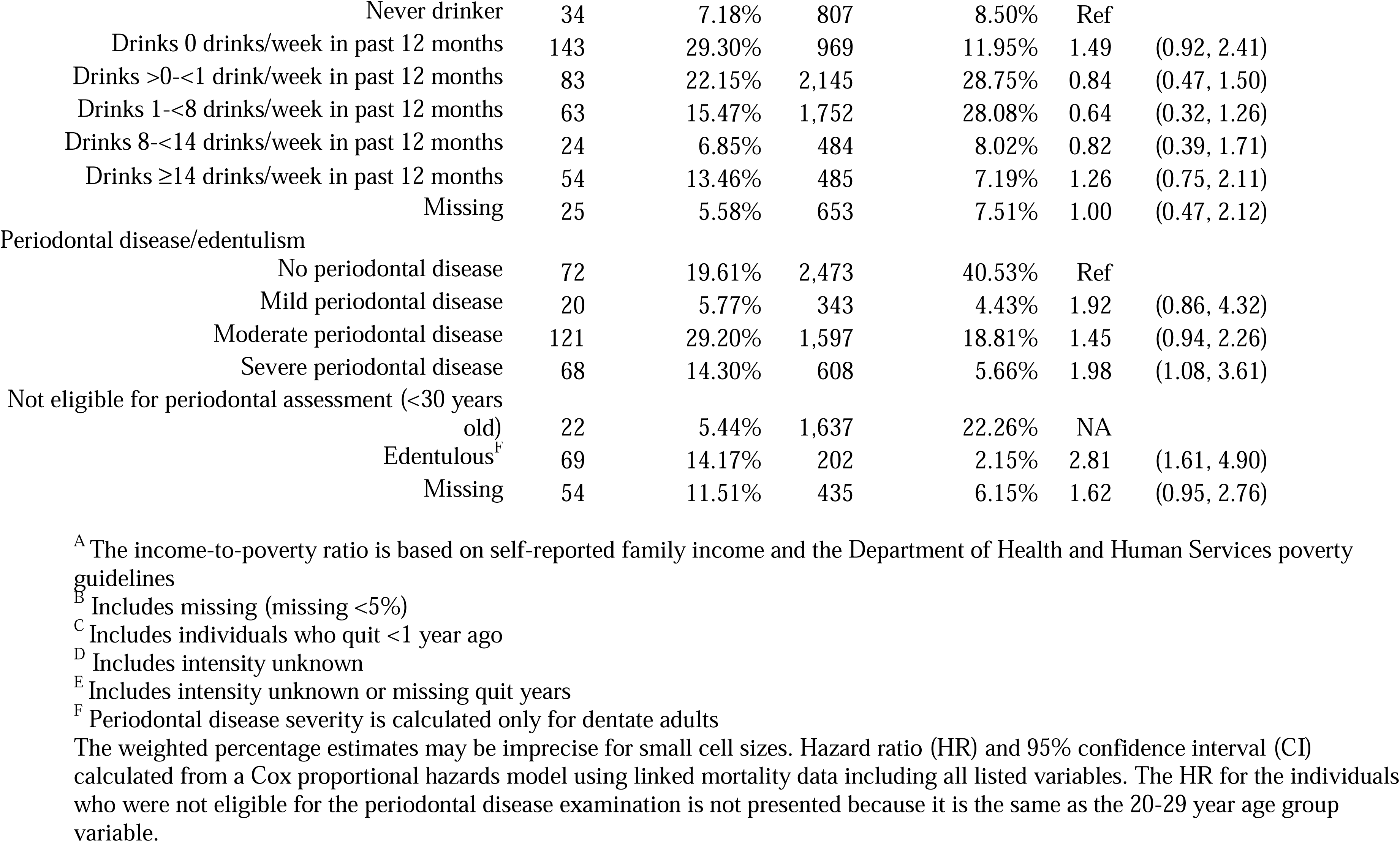
Selected characteristics of participants and associations with overall mortality in the linked oral microbiome and mortality NHANES files 2009-2012.

All continuous alpha diversity metrics were inversely associated with all-cause mortality after minimal adjustment (e.g., HR_Observed ASVs_ per SD=0.82; 95% CI=0.68-0.98), but associations were attenuated after full adjustment (Figure 1 and Supplemental Table 1). In fully adjusted models, only the Shannon-Weiner index association remained statistically significant (HR per SD=0.85; 95% CI=0.74-0.98). The associations for quartiles of alpha diversity were inverse, but only the association for the highest quartile for the Simpson index reached statistical significance (HR_Q4_=0.62; 95% CI=0.39-0.99; Supplemental Table 1). For beta diversity, PCoA2 from the Bray-Curtis dissimilarity (HR per SD=0.83; 95% CI=0.73-0.93) and PCoA1 from weighted UniFrac (HR per SD=0.86; 95% CI=0.75-0.98) matrices were statistically significantly associated with all-cause mortality after full adjustment (Figure 1 and Supplemental Table 1).

**Figure 1.**
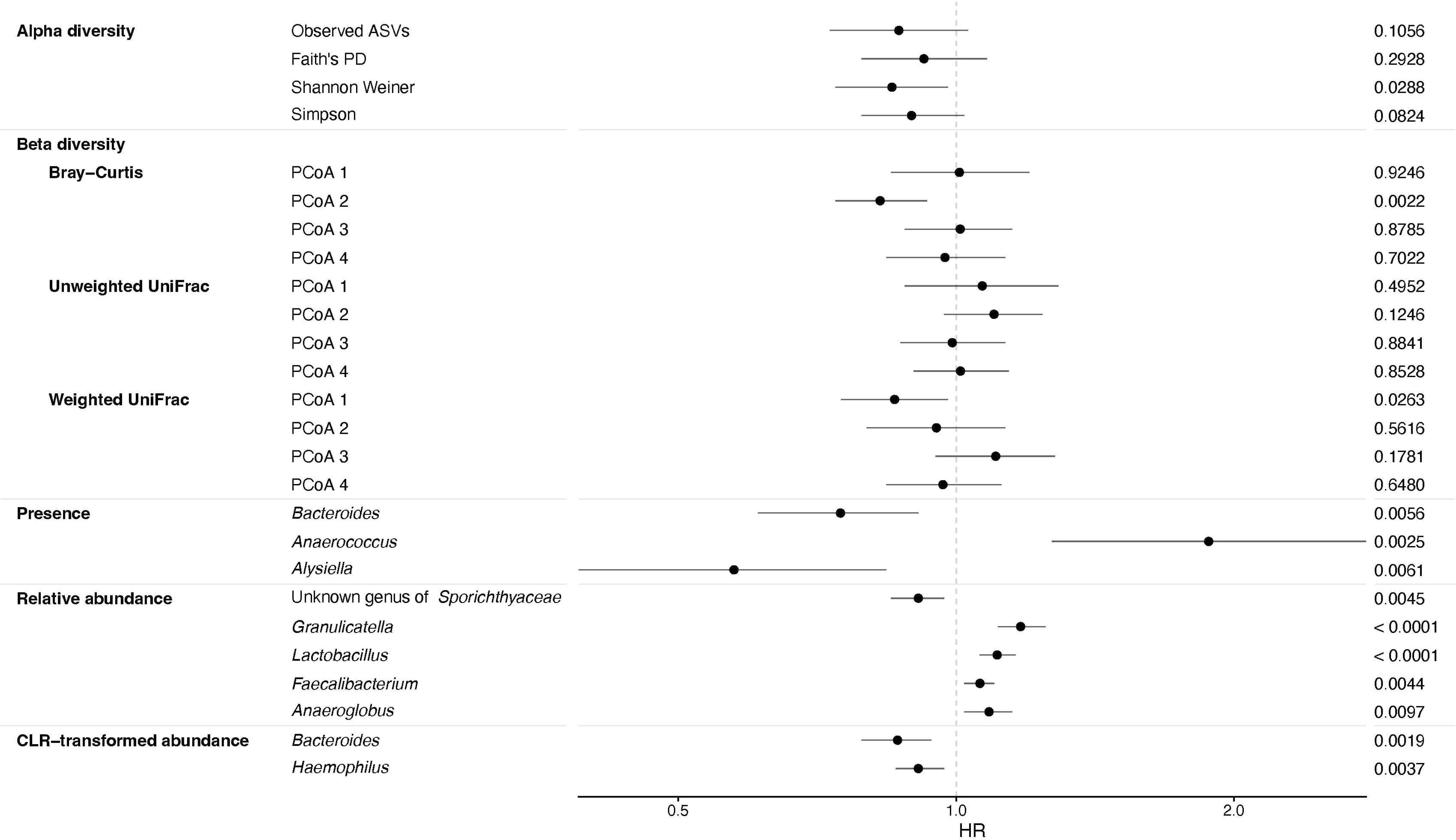
Forest plot of the estimated hazard ratios (HR) with 95% confidence intervals and p-values for the associations of alpha diversity (i.e., observed amplicon sequence variants [ASV], Faith’s Phylogenetic Diversity [PD], Shannon Weiner, and Simpson indices), beta diversity (i.e., Bray-Curtis, unweighted UniFrac, and weighted UniFrac) principal coordinate analysis (PCoA) vectors, and specific genera with all-cause mortality in the linked oral microbiome and mortality NHANES files 2009-2012. The Cox proportional hazards models included adjustment for age, age splines, sex, self-reported race and ethnicity as categorized by NHANES, education, marital status, income-to-poverty ratio, body mass index category, detailed smoking history, alcohol consumption, periodontal disease/edentulism, prevalent diabetes or hypertension, and use of prescription medications for antibiotics, anti-lipidemic drugs, inhaled respiratory drugs, and drugs for treatment of gastroesophageal reflux disease. Alpha diversity, beta diversity, relative abundance, and centered log-ratio (CLR) transformed abundance associations were modeled as the per standard deviation change in the estimate. For the presence analysis, no genera were significantly associated with all-cause mortality at a Bonferroni adjusted p-value (p-value < 0.05/133 = 0.0004), but genera with a p-value less than 0.01 are presented. All of the calculated genus-level presence associations are included in Supplemental Table 2. For the relative abundance analysis, two genera were statistically significant at the Bonferroni adjusted p-value (p-value < 0.05/103 = 0.0005) and genera with a p-value less than 0.01 are presented. All of the calculated genus-level relative abundance associations are included in Supplemental Table 3. For the CLR-transformed abundance analysis, no genera were significantly associated with all-cause mortality at a Bonferroni adjusted p-value (p-value < 0.05/103 = 0.0005), but genera with a p-value less than 0.01 are presented. All of the calculated genus-level CLR-transformed abundance associations are included in Supplemental Table 4.

After Bonferroni correction, none of the genus-level presence associations were statistically significant in fully adjusted models, although some strong associations were noted (Figure 1 and Supplemental Table 2). For relative abundance, each SD increase in *Granulicatella* or *Lactobacillus* was associated with 1.17 (95% CI=1.11-1.24) and 1.11 (95% CI=1.06-1.16) times the mortality risk, respectively, which remained statistically significant after Bonferroni correction (Figure 1 and Supplemental Table 3). However, none of the CLR-transformed abundances were statistically significant in fully adjusted models, although the association with *Lactobacillus* was similar to the relative abundance analysis (HR 1.13; 95% CI: 0.99-1.28; Figure 1 and Supplemental Table 4). In the combined presence and relative abundance analysis, compared to participants with no *Bacteroides* detected, participants in the highest tertile had reduced mortality risk (HR=0.54; 95% CI=0.40-0.74; p_trend_=0.0001; Figure 2 and Supplemental Table 5).

**Figure 2.**
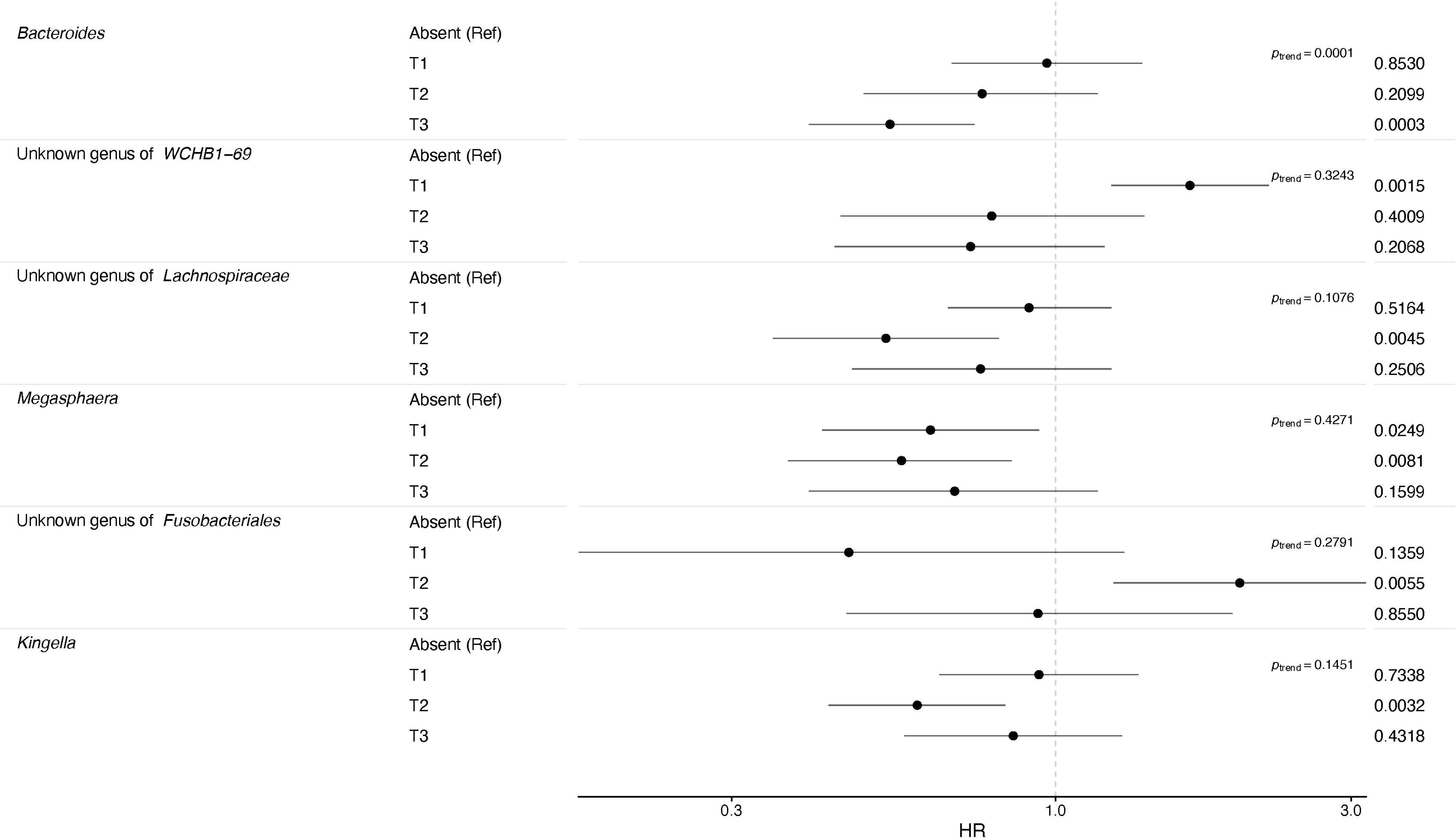
Forest plot of the estimated hazard ratios (HR) with 95% confidence intervals and p-values for the associations of the combined genus-level presence and relative abundance with all-cause mortality in the oral microbiome and linked mortality NHANES files 2009-2012. The Cox proportional hazards models included adjustment for age, age splines, sex, self-reported race and ethnicity as categorized by NHANES, education, marital status, income-to-poverty ratio, body mass index category, detailed smoking history, alcohol consumption, periodontal disease/edentulism, prevalent diabetes or hypertension, and use of antibiotics, anti-lipidemic drugs, inhaled respiratory drugs, and prescription drugs for treatment of gastroesophageal reflux disease. The reference value is the absence of the specific genus compared to the calculated tertiles of relative abundance for that genus. For the p-value of the trend, only *Bacteroides* with a p-trend of 0.0001 was statistically significant at the Bonferroni adjusted p-value (p-value < 0.05/83 = 0.0006), but genera with a p-value less than 0.01 for the association with any tertile are presented. All of the calculated combined genus-level associations are included in Supplemental Table 5.

## Discussion

We found some oral microbial population characteristics to be independently associated with all-cause mortality in this large, representative cohort of the 20- to 69-year-old US population. The alpha diversity measures incorporating richness and evenness (i.e., the Shannon and Simpson indices) were significantly inversely associated with mortality. Some significant associations were also observed with the beta diversity matrices which incorporate the relative abundance of the ASVs (i.e., Bray-Curtis and weighted UniFrac) which suggests that the oral microbial community itself may be related to health and disease. At the genus-level, a higher relative abundance of *Granulicatella* and *Lactobacillus* were associated with increased all-cause mortality, while compared with individuals without detectable *Bacteroides*, a high relative abundance of *Bacteroides* was associated with decreased all-cause mortality. These three genera are common oral species, but *Granulicatella* has been associated with endocarditis and bacteremia,^13^ *Lactobacillus* has been associated with increased risk of lung cancer,^4^ and *Bacteroides* has been implicated in periodontal disease^1^ and health.^14^

This study has important strengths as it is representative of the adult US population and prospective in nature with relatively long follow-up from NDI linkage. We included thorough adjustment for many confounders, particularly measured periodontal disease status and edentulism, which provides strong evidence for independent microbiome associations. However, there are some limitations to note. Due to the use of 16S rRNA gene sequencing, we were only able to accurately assign the bacterial taxa to the genus-level. The use of shotgun metagenomics in future studies would allow for more granular and imputed functional associations. There were also relatively few deaths over the follow-up, which precluded the study of cause-specific mortality outcomes. Finally, there is the possibility for residual confounding from unmeasured or uncontrolled factors.

In conclusion, some measures of the oral microbiome were independently associated with all-cause mortality in a nationally representative US cohort. Our findings highlight the importance of the oral microbiome and oral health to overall human health and more research could lead to a greater understanding of these associations.

## Data Availability

Unlinked public-use National Health and Nutrition Examination Survey (NHANES) data files are available on the NHANES website (https://www.cdc.gov/nchs/nhanes). The linked mortality (National Death Index) files are available in a public-use version, available online (https://www.cdc.gov/nchs/data-linkage/mortality.htm), and a restricted-use file available for analysis at the National Center for Health Statistics Research Data Center (https://www.cdc.gov/rdc/index.htm). The restricted-use mortality file was used for this analysis.

## Acknowledgements

We would like to acknowledge Gail Ackermann, Amnon Amir, James Gaffney, Grant Gogul, Greg Humphrey, Tara Schwartz, and Weihong Xiao for their role in project management and data production. This study was supported in part by a grant from the National Institute for Dental and Craniofacial Research (R01DE023175), a grant from the US Food and Drug Administration’s Center for Tobacco Products, a grant from the National Cancer Institute Informatics Technology for Cancer Research (1U24CA248454-01), the Intramural Research Program of the National Cancer Institute, and Federal funds from the National Cancer Institute, National Institutes of Health, under NCI Contract number 75N910D00024. We also acknowledge the research contributions of the Cancer Genomics Research Laboratory for their expertise, execution, and support of this research in the bioinformatics analysis of generated data. The content of this publication are those of the authors and does not necessarily reflect the views or policies of the Department of Health and Human Services or the official position of the Centers for Disease Control and Prevention. The mention of trade names, commercial products, or organizations does not imply endorsement by the US Government.

